# LUNG ULTRASOUND FINDINGS IN COVID-19 RESPIRATORY DISEASE AND CORRELATION TO DISEASE SEVERITY

**DOI:** 10.1101/2020.09.28.20182626

**Authors:** Julia Burkert, Hannah Dunlop, Rachel Stewart, Adam Treacy, Robert Jarman, Paramjeet Deol

## Abstract

Lung ultrasonography has emerged as a promising imaging modality during the COVID-19 pandemic with potential use in triage, diagnosis, prognosis and disease progression.

This retrospective observational cohort study carried out in a busy urban Emergency Department in the United Kingdom provides a systematic analysis of lung ultrasound findings (pleural irregularities, B-lines, consolidations, pleural effusions) in SARS-CoV-2 positive patients and correlates these findings to disease severity as defined by oxygen-deficiency based on the Berlin Criteria for ARDS.

Our results show that wide B-lines, as well pleural irregularities and subpleural consolidations are frequent findings in COVID-19 disease. Lung abnormalities occur bilaterally, interspersed with normal lung, and without significant predilection for specific lung areas. Wide B-lines are a strong feature in COVID-19 infection. We also describe a finding of small localised peri-pleural effusions in 8.3% (95% C.I. 5.9-10.8%) of lung zones.

Disease severity correlates strongly to the frequency of abnormal ultrasound findings. Irregular pleura and subpleural consolidations increase from 40% (95% C.I. 33.3-46.1%) and 27.7% (C.I.95% 21.8-33.5%) of zones affected in mild disease to 85.7% (C.I.95% 79.8-91.7%) and 66.2% (C.I.95% 58.1-74.2%) in severe disease. Wide B-lines increase from 15.6% (C.I.95% 10.9-20.4%) to 45.1% (C.I.95% 36.7-53.6%). There is an inverse correlation to the amount of normal lung seen, decreasing from 57.1% (C.I.95% 50.6-63.6%) to 6% (C.I.95% 2.0-10.1%) of lung zones.

These results contribute to a more thorough understanding of the lung changes in COVID-19 pneumonia and enhance the evidence base for the application of ultrasound in triage, diagnosis, treatment, and prognosis in SARS-CoV-2 infection.

## INTRODUCTION

COVID-19 is the disease caused by SARS-CoV-2 infection, first identified in China in 2019, which spread rapidly across the world with pandemic status declared in March 2020 by the World Health Organisation(1). Clinically the infection has a broad range of manifestations, but presents predominantly as a respiratory illness with a wide range of severity, including asymptomatic cases, mild flu-like symptoms, as well as viral pneumonia, and in extreme cases ARDS-like illness with a high mortality rate(2).

Evidence from computerised tomography (CT) imaging of the chest showed that COVID-19 respiratory disease gives rise to a classic appearance of patchy peripheral ground-glass opacities (GGO) with ill-defined margins and inter- and intralobular septal thickening leading to so-called “crazy paving” cobble-stone patterns. Areas of ground glass opacification and small consolidations occur bilaterally, peripherally and all lung segments can be involved. There are also associated pleural abnormalities(3)(4)(5). Whilst CT is the imaging gold-standard for diagnosis and assessment of disease severity in COVID-19 pneumonia, it has significant disadvantages which become particularly poignant in the setting of a pandemic. These include radiation exposure which has to be considered particularly in paediatric and pregnant patients, infection control problems, the need for specialist reporting and the length of time needed from scan to report.

Since the start if the COVID-19 outbreak, lung ultrasound has attracted significant attention with a large amount of literature dedicated to its potential use and benefits, especially given the relatively poor sensitivity and specificity of CXR for the changes typical for COVID-19(6). The usefulness of lung ultrasound in the context of viral pneumonias has been previously explored in the H1N1 (swine-flu) and H7N9 (avian influenza) pandemics in 2009 and 2013 respectively(7)(8).

The typical findings seen on lung ultrasound of Covid-19 pneumonia include pleural irregularities, subpleural consolidations, as well as vertical artefacts known as B-lines, which signify interstitial abnormalities. B-lines appear in a variety of morphologies, from thin scattered lines to wide confluent bands that can span an entire rib space. Large consolidations and pleural effusions are rare(9)(10)(11)(12)(13)(14)(15). These abnormalities are found in patchy distribution throughout both lungs and are similar to those described for ARDS. The changes appear to relate to disease severity with anterior involvement a high predictor of need for respiratory support (16). Although sensitivity of lung ultrasound for detection of COVID-19 lung disease is comparable to CT with 90-94%, its poor specificity, shown to be ranging between 7-83% with a high dependence on disease prevalence has cast doubt on its usefulness (16)(17)(18)(19).

Many potential applications have been suggested for the use of lung ultrasound in this pandemic ranging from triage to diagnosis, disease monitoring and therapeutic guidance, however none of these applications have found standardisation and validation as of yet. Despite growing evidence, methodologically sound and detailed studies describing lung ultrasound findings in COVID-19 are still evolving. Differences in scanning approaches, disease severity of population scanned, abnormalities examined, and small sample sizes have led to significant heterogeneity in findings, particularly with respect to irregular pleura and subpleural consolidations which have been described in ranges from 13% to 100% of patients(20)(21)(14)(22)(23)(24)(25)(26)(12)(27). With respect to examining sensitivity and specificity of lung ultrasound, no consensus definition is yet reached as to what constitutes a scan ‘positive’ for COVID-19 infection(28). In order to characterise and validate the lung ultrasound features in COVID-19 pneumonia with a view to finding potential abnormality patterns that may increase specificity, more systematically analysed data is needed

In this retrospective observational cohort study, a 12-zone scanning approach is used to assess each lung zone for 8 different lung findings (pleural irregularities, B-lines in 3 different morphologies, large and small consolidations, large and small pleural effusions). The percentages of total lung zones affected by each abnormality are compared between different anatomical locations within the lung, and the findings are also correlated to clinical severity as determined by SpO_2_/FiO_2_ (S/F) ratio, with cut-off values based on the Berlin Criteria for the diagnosis of ARDS(29). The findings from our study could further guide future research into the utility of lung ultrasound in the diagnosis and prognostication of COVID-19.

## ETHICS

This study was approved by the Health Research Authority (HRA) (no.286642).

## METHODS

### Selection of patients

Patients who had presented to the ED in a 6-week period (from 15. March to the 30. of April 2020) with symptoms consistent with COVID-19(30) and also had received a lung ultrasound by one of the ED doctors involved in the study as part of their routine clinical assessment were considered for the study (n=60). Retrospectively, SARS-CoV-2 infection status as determined by RT-PCR was ascertained by cross-referencing patient’s notes on *e-Record™ patient*. This resulted in 44 patients.

Patients were divided into three clinical severity groups based on oxygenation deficit as determined by SpO_2_/FiO_2_ (S/F) ratio: mild (S/F>315), moderate (S/F 148-315), and severe (S/F <148). The cut-off values were based on the Berlin Diagnostic criteria for ARDS (29) and were determined using an equivalence calculation for translating PaO_2_/FiO_2_ (P/F) ratios into S/F ratios as suggested by Rice *et al* (31)(32) (Table 2).

**Table 1.**
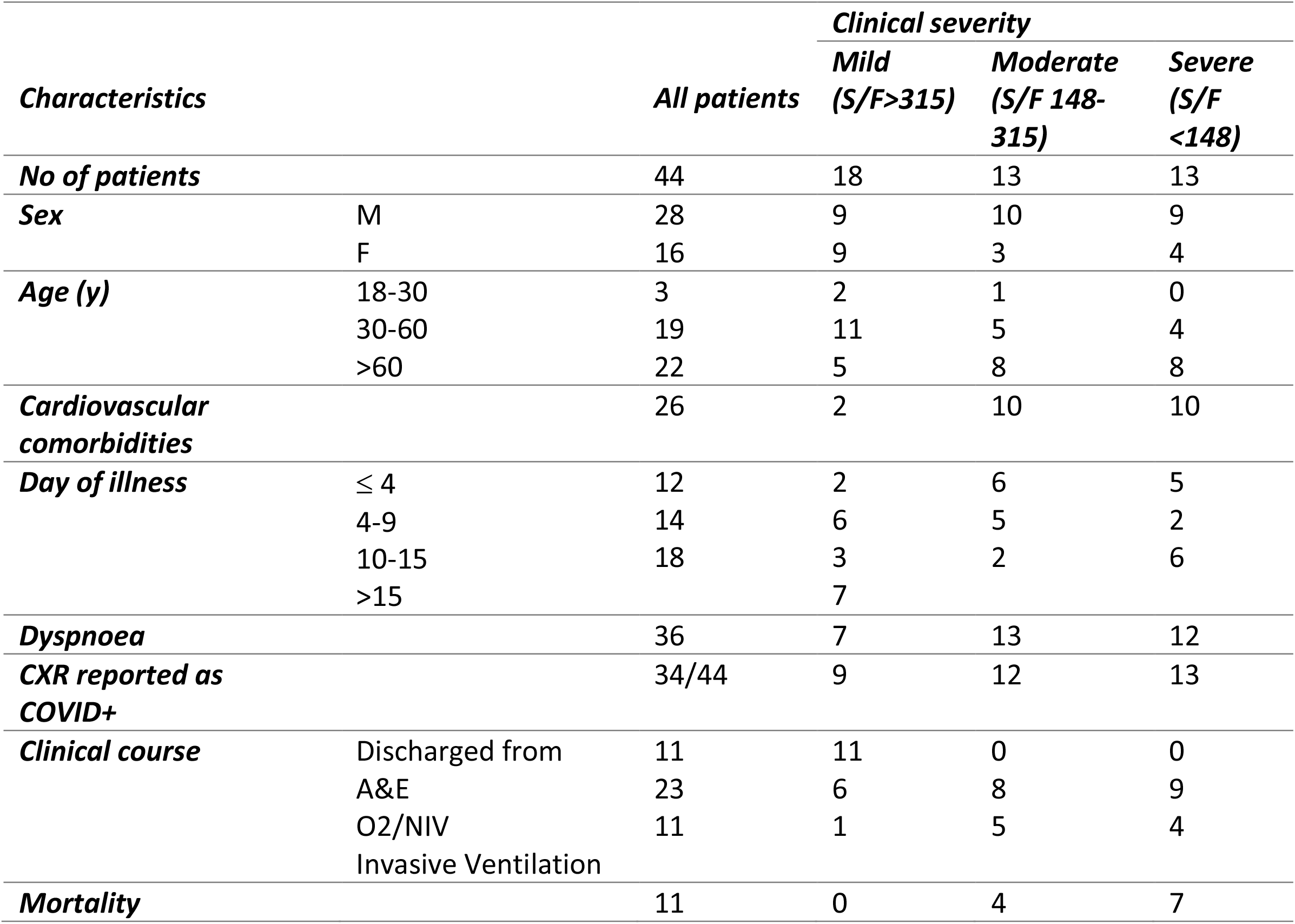
Patient Characteristics

**Table 2.**
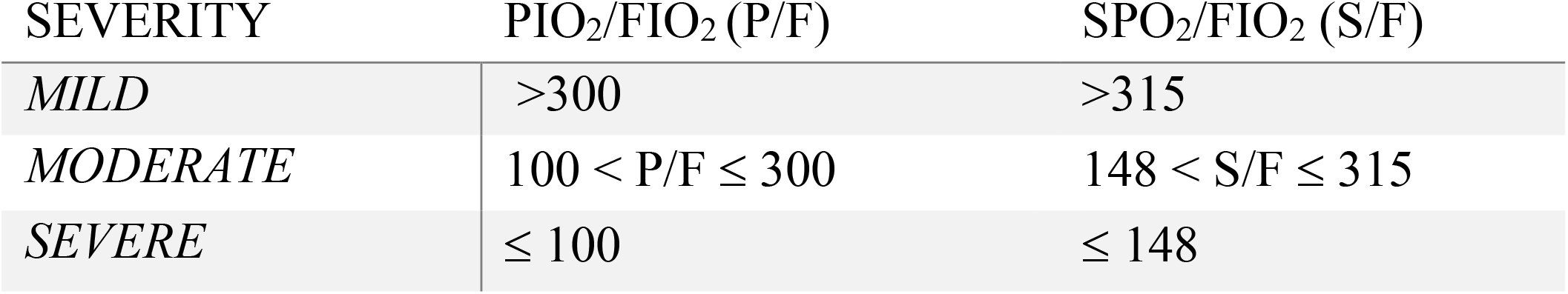
disease severity grouped into 3 categories according to SpO2/FiO2 ratios and cut-off values based on the Berlin Criteria for ARDS. Left column showing equivalence conversion to PaO2/FiO2 ratios.

### Ultrasound

Lung ultrasound scans were performed by one of three doctors in the Emergency Department, one of which was an expert with >5 years experience in POCUS and both other doctors had completed a formal ultrasound fellowship, as well as ten supervised 12-zone LUNG ULTRASOUND scans prior to the study. Two ultrasound machines were used in the department - a SonoSite Xport ™ (FUJIFILM SonoSite Inc, Bothell, WA, USA) with set to abdominal pre-set with Tissue Harmonic Imaging (THI) and Compound Imaging (CI) manually turned off, or a GE Venue™ (GE Healthcare, Chicago, Illinois, US) with lung pre-set. The intercostal spaces were scanned sagitally with a low-frequency transducer (3-5MHz) at a depth of 13-16 cm. The focal zone was set to the pleural line. Each lung zone was scanned following a ‘lawn-mower approach’ and the image with most abnormalities was saved. Still images and clips of each zone were saved for analysis.

Twelve lung zones were assessed where the patients’ clinical condition allowed. Zones were identified as follows: each hemithorax was divided by anterior and posterior axillary lines into anterior, lateral and posterior areas, which were further subdivided into superior and inferior zones, resulting in 6 scanning zones per hemithorax. Labelling was set according to laterality of hemithorax, R (right) and L (left)), and zones were numbered as 1 and 2 for superior and inferior anterior zones, 3 and 4 for superior and inferior lateral zones, and 5 and 6 for superior and inferior posterior zones (Figure 1).

**Figure 1.**
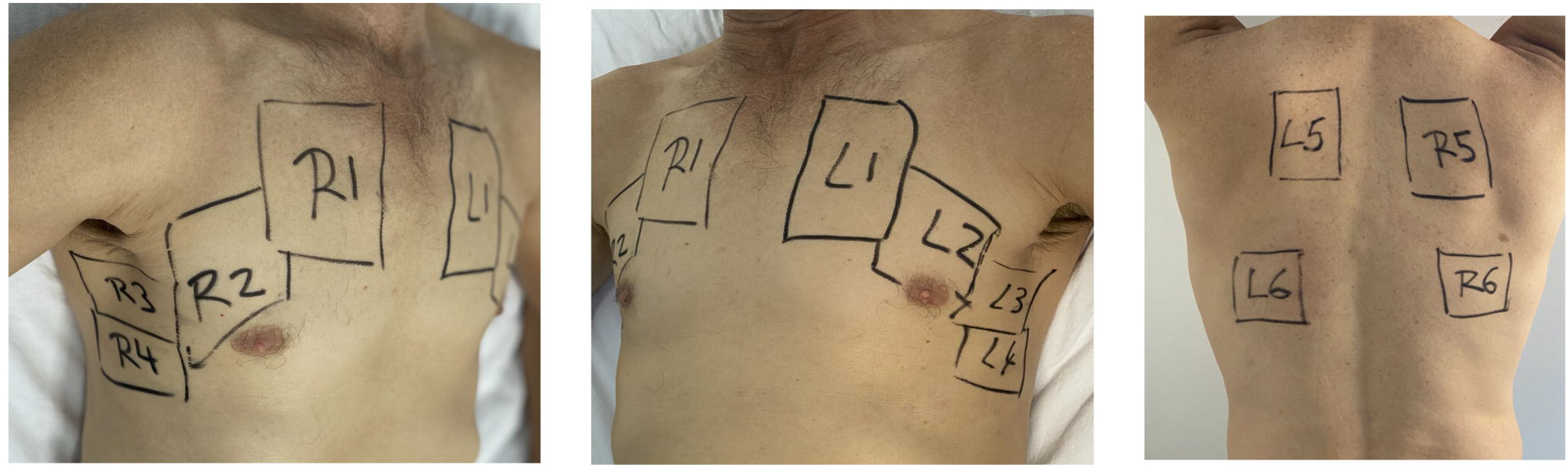
Lung Zones scanned for assessment of COVID-19 related changes. Each hemithorax was divided into 6 Zones as depicted. Anterior, lateral and posterior determined as separated by anterior and posterior axillary lines and subdivided into superior and inferior zones.

Machines and transducers were doubly disinfected with between patients and protective hygiene precautions were taken in form of full PPE according to local hospital guidelines.

### Review process

Retrospectively, all images and videoclips from SARS-CoV-2 positive patients were anonymised and reported independently by two of the ED clinicians trained in ultrasound taking part in the study. Reporting was carried out using an ultrasound proforma that was specifically developed for assessment of COVID-19 lung ultrasound scans in the department (Figure 2). All abnormalities seen within an intercostal space were taken into account. Reports were compared for agreement and categorised as ‘agree’ or ‘disagree. In cases of disagreement, a third doctor with experience in interpreting lung ultrasound scans was consulted and the consensus result was used. The reports were compared categorically, and inter-rater reliability was assessed using Cohen’s Kappa.

**Figure 2.**
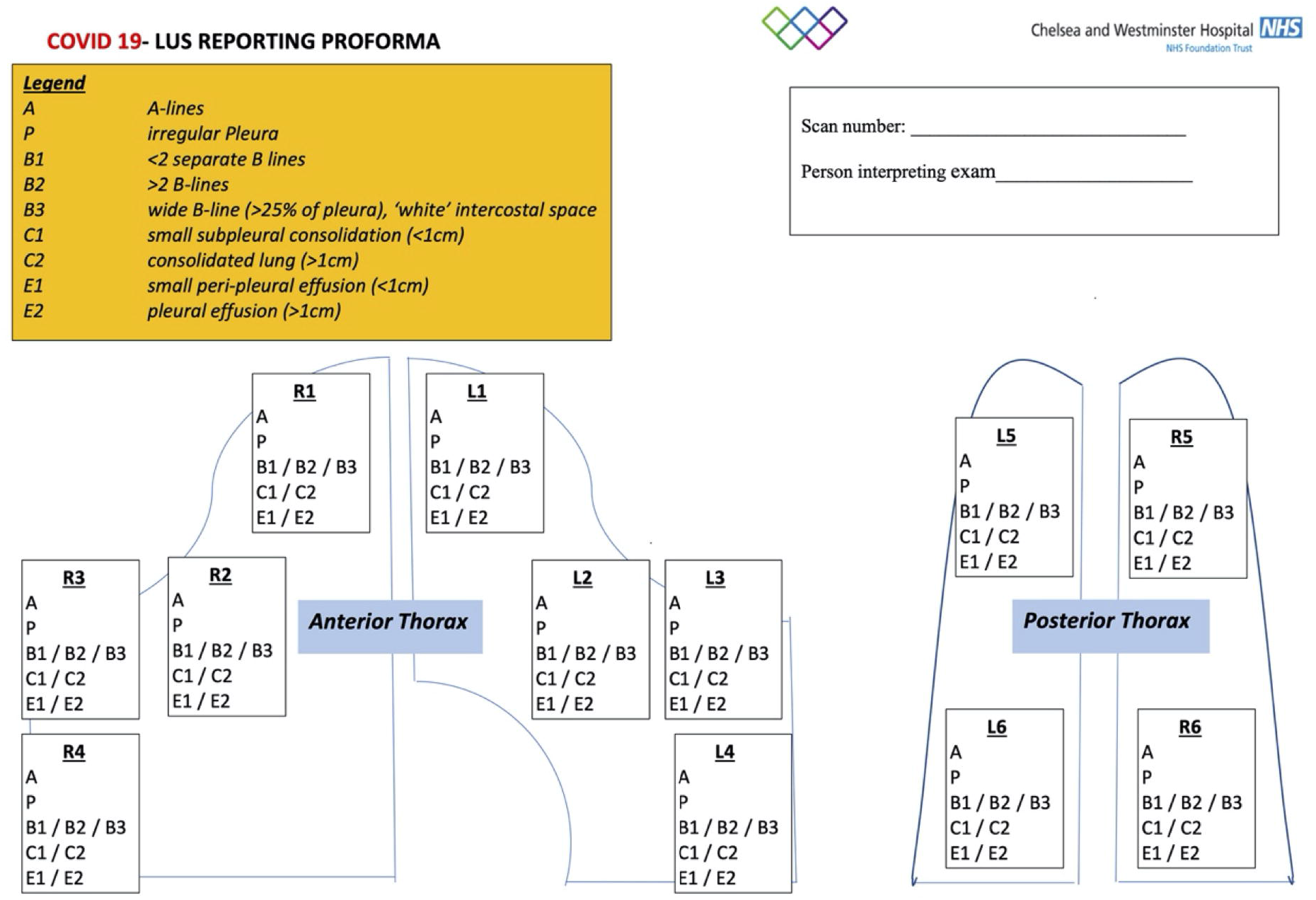
Reporting proforma used for the analysis of lung ultrasound scans

The sonographic clips or images were scored for the presence of the following ultrasound appearances: A-line pattern (A), pleural irregularity (P), small peripheral consolidation (<1cm) (C1), larger consolidation (C2), B-lines: ≤ 2 isolated (B1), >2 B-lines (B2), thick B-lines occupying >25% of the pleura at origin or confluent B-lines occupying the entire intercostal space (B3), and pleural effusions: small (<1cm) (E1), large (E2) (Figure 2).

### Statistical analysis

Frequencies of lung ultrasound findings, as well as 95% confidence intervals were calculated for each lung ultrasound finding across all scans, as well as across severity categories and different anatomical lung areas. Results were compared between upper (zones 1&5), middle (zones 2&3), and lower (zones 4&6) lung areas, as well as between anterior (zones 1&2), lateral (zones 3 &4), and posterior (zones 5&6), as well as between mild, moderate and severe disease severities. Pearson’s chi-square tests with 3×2 contingency tables were performed to ascertain statistically significant differences. Desired p-value thresholds of 0.05 were adjusted using Bonferroni correction to allow for multiple comparisons. To establish strength of correlation between lung abnormalities and disease severity, linear regression analysis was performed with calculation of adjusted Pearson’s correlation coefficients (r). Statistical analyses were carried out using MS Excel [Version16.41] Microsoft Corporation, Redmond, WA, USA.

## RESULTS

Forty-four patients out of sixty patients who had an initial 12-zone lung ultrasound performed in the ED as part of their initial clinical assessment were found to be positive for SARS-CoV-2 infection and included in the study. Patient characteristics are shown in Table 1.

The patient demographics included 28 males and 16 females, 22 patients over the age of 60 and 26 having prior cardiovascular morbidities. Dyspnoea was the main presenting complaint in 36 patients. Patients were grouped into severity groups according to S/F ratios as described in Table 2, resulting in 18 patients classified as mild, 13 patients as moderate and 13 as severe disease.

Where possible, 12-zone ultrasound scans were performed, however in some patients, clinical severity did not allow for repositioning to access all 12 zones. This resulted in scans of a total of 492 zones which were anonymised and retrospectively independently reported by two physicians. the inter-rater reliability agreement was 91% with a Cohen’s K value of 0.8.

### Frequency of different lung abnormalities

All of the 492 analysed zones were independently assessed for the presence of eight different ultrasound findings. Overall frequencies and 95% Confidence Intervals (C.I.) were calculated (Table 3). Overall, the most prevalent findings were irregular pleura (P) and small subpleural consolidations of less than 1cm depth (C1), found in 59.6% (95% C.I. 25.1-33.1%) and 44.9% (C.I.95% 40.5-49.3%) of all scans respectively. Wide confluent B-lines, spanning the entire rib space were also seen in a high percentage of scans (30.1%, 95% C.I. 26.0-34%). Other B-line morphologies were seen to a lesser extent. Notably, we also demonstrated a proportion of small peri-pleural effusions (E1) in 8.3% (C.I.95% 5.9-10.8%) of all scans. Large consolidations and pleural effusions were seen infrequently comprising less than 2.6% of all scans (Figure 3). Each lung abnormality was found in isolation, as well as together with other abnormalities described above. Abnormal lung was frequently interspersed with areas of normal lung with A-line pattern. The abnormalities were found throughout all lung zones and occurred bilaterally (Figures 4A-C).

**Table 3.** overall frequency of lung ultrasound findings and 95% confidence intervals amongst all lung zones analysed

|  | **Positive (P) (%)** | **95% C.I. (%)** |
| --- | --- | --- |
| **A** | 29.1 | 25.1-33.1 |
| **P** | 59.6 | 55.2-63.9 |
| **B1** | 14.8 | 11.7-18.0 |
| **B2** | 2.9 | 1.4-4.3 |
| **B3** | 30.1 | 26.0-34.1 |
| **C1** | 44.9 | 40.5-49.3 |
| **C2** | 2.6 | 1.2-4.1 |
| **E1** | 8.3 | 5.9-10.8 |
| **E2** | 2.2 | 0.9-3.5 |

**Figure 3.**
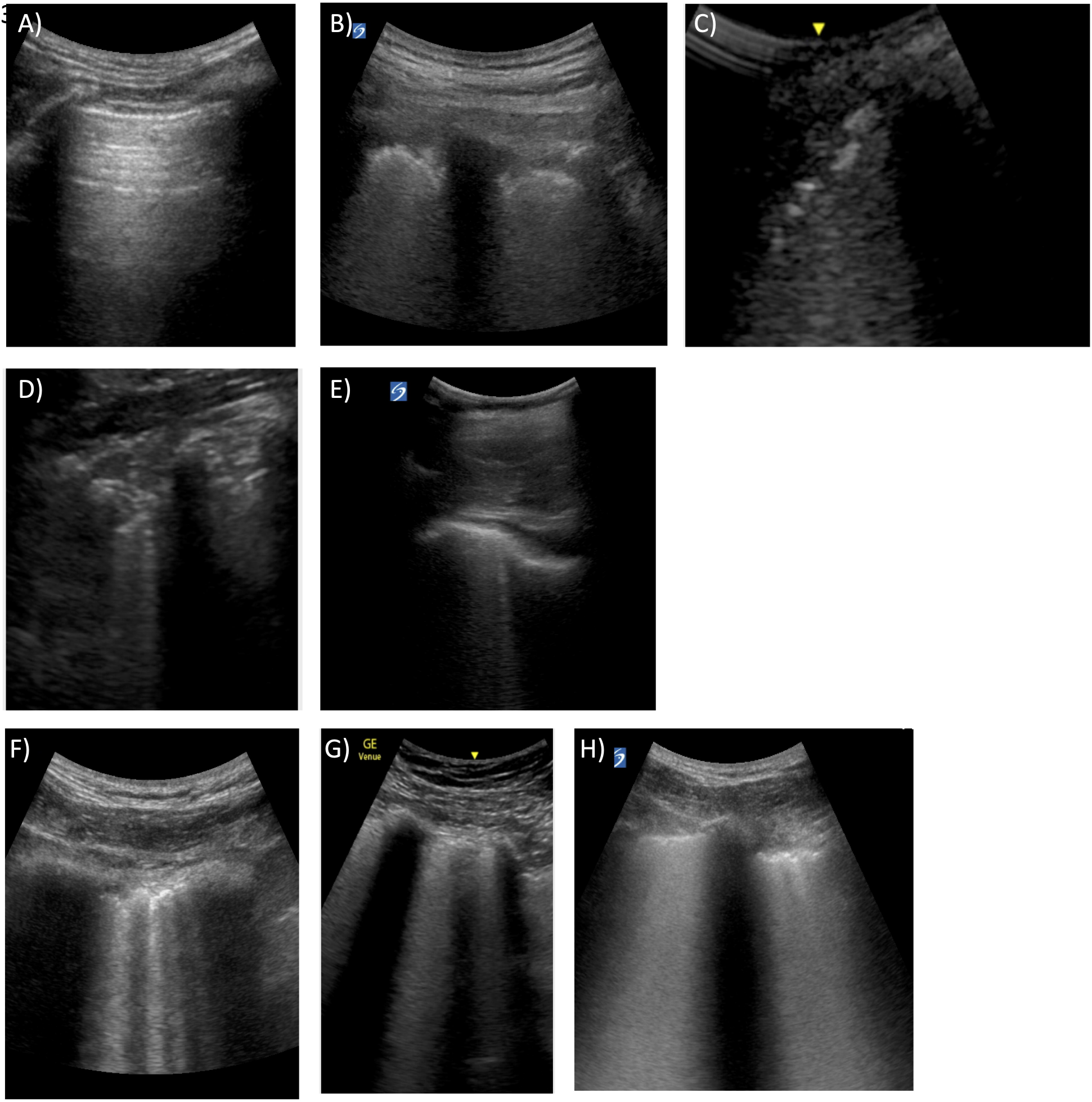
Images of typical lung ultrasound appearances in COVID-19 respiratory disease. A) A-lines indicating normal lung, B) irregular and thickened peura, C) irregular and disrupted pleura, D) small subpleural consolidations (< 1cm), E) small localised effusions, F) separate B-lines, G) broad band-like B-lines, H) coalesced B-lines occupying the entire rib space.

**Figure 4.**
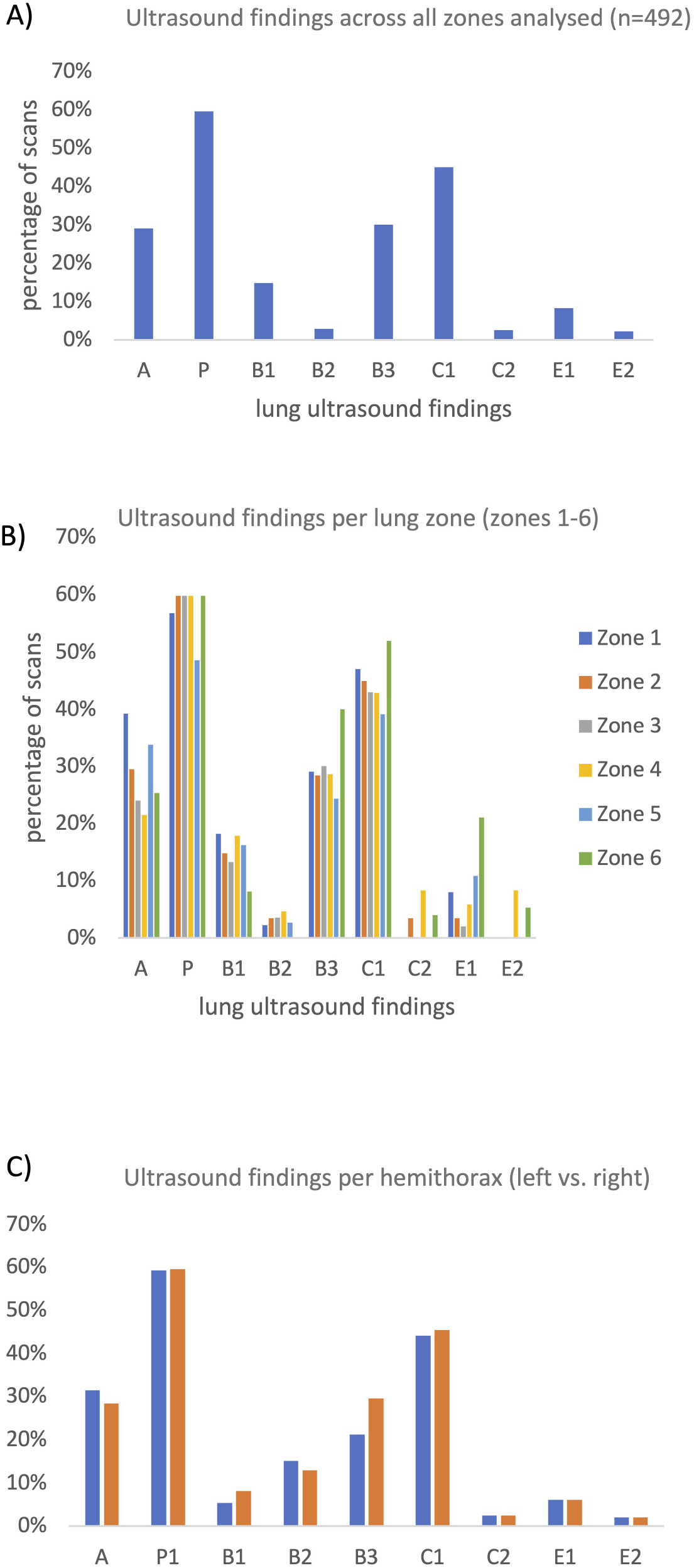
Bar graphs showing the percentage of lung zones affected by each abnormality as seen on lung ultrasound. A) percentage of abnormalities amongst all zones analysed (n = 492). B) percentage of positive findings for each of the 6 lung zones. C) percentage of positive findings comparing left and right hemithorax.

### Anatomical distribution of lung abnormalities

Different areas of the lung were analysed for differences in distribution of the lung abnormalities (Table 4). When comparing transverse planes of the lung, i.e. apical (zones 1&5) vs. middle (zones 2&3) vs. basal zones (zones 4&6). Small peripleural effusions (E1) were found more commonly in the apical and basal lung zones, seen in 9.3% (C.I.95% 4.9-13.7%) and 13.2% (C.I.95% 7.9-18.5%) of scans, compared to the middle with 2.9% (C.I.95% 0.4-5.5%).

**Table 4.**
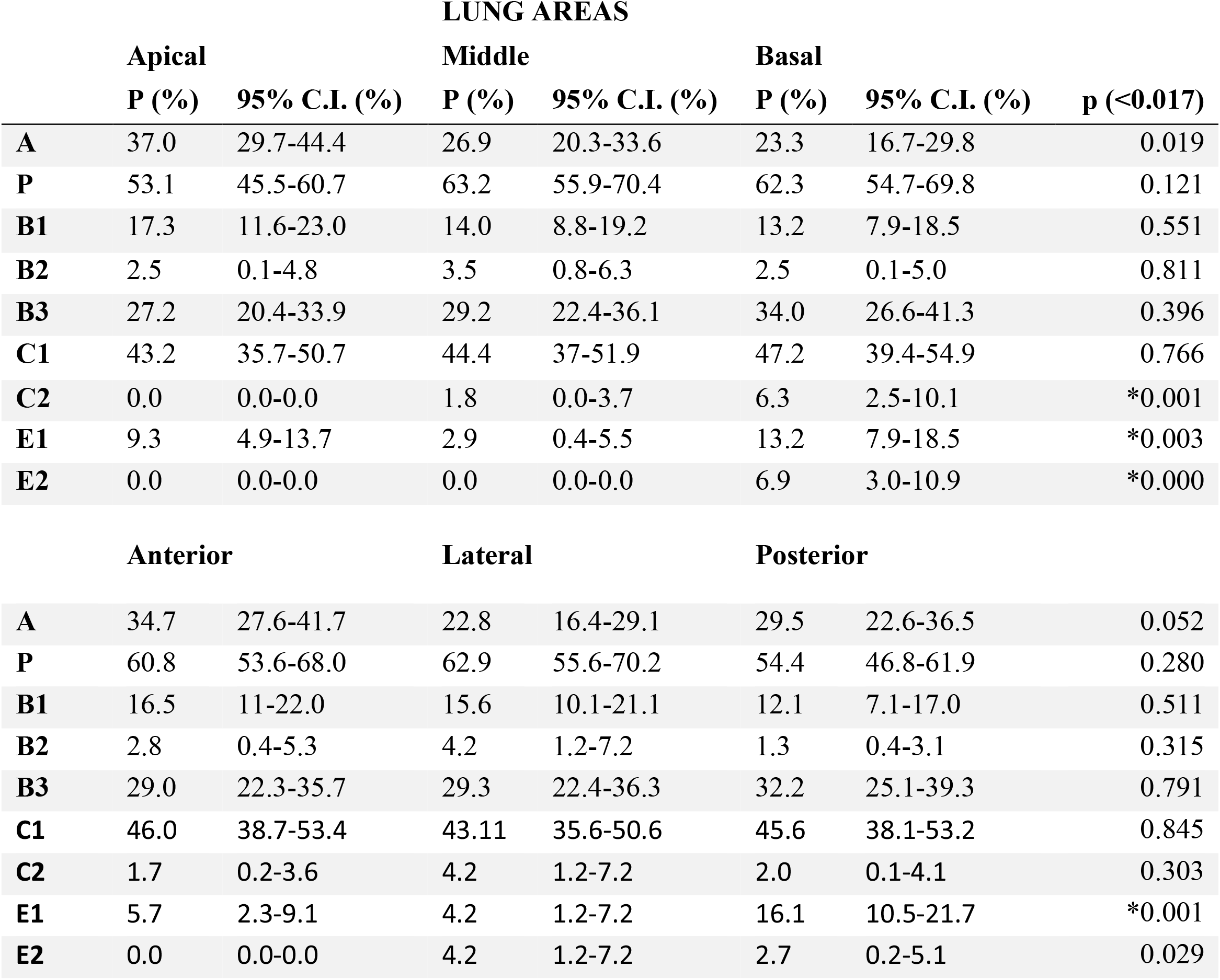
Percentages of lung ultrasound findings and 95% confidence intervals separated by lung areas. The desired p-value was reached by Bonferroni correction to allow for multiple comparisons. Asteriks indicate statistical significance.

When comparing coronal planes of the lung, i.e. anterior (zones 1&2) vs. lateral (zones 2&3) vs. posterior zones (zones 5&6), small peripleural effusions were seen to a significantly higher extent in the posterior zones (16.1%, C.I.95% 0.5-21.7%), compared to anterior and lateral zones showing the finding in 5.7% (C.I.95% 2.3-9.1%) and 4.2% (C.I.95% 1.2-7.2%). Gravity-dependent findings such as large pleural effusions (E2) were only seen in basal and posterolateral zones, found in 6.9% of scans from those zones. Large consolidations were found to a similar extent in coronal sections of the lung, but showed predilection for basal zones, occurring in 6.3% (C.I.95% 2.5-10.1%) of basal scans.

All other lung findings that were examined did not display significant differences between lung zones both, in transverse and coronal planes (Figure 5A,B).

**Figure 5.**
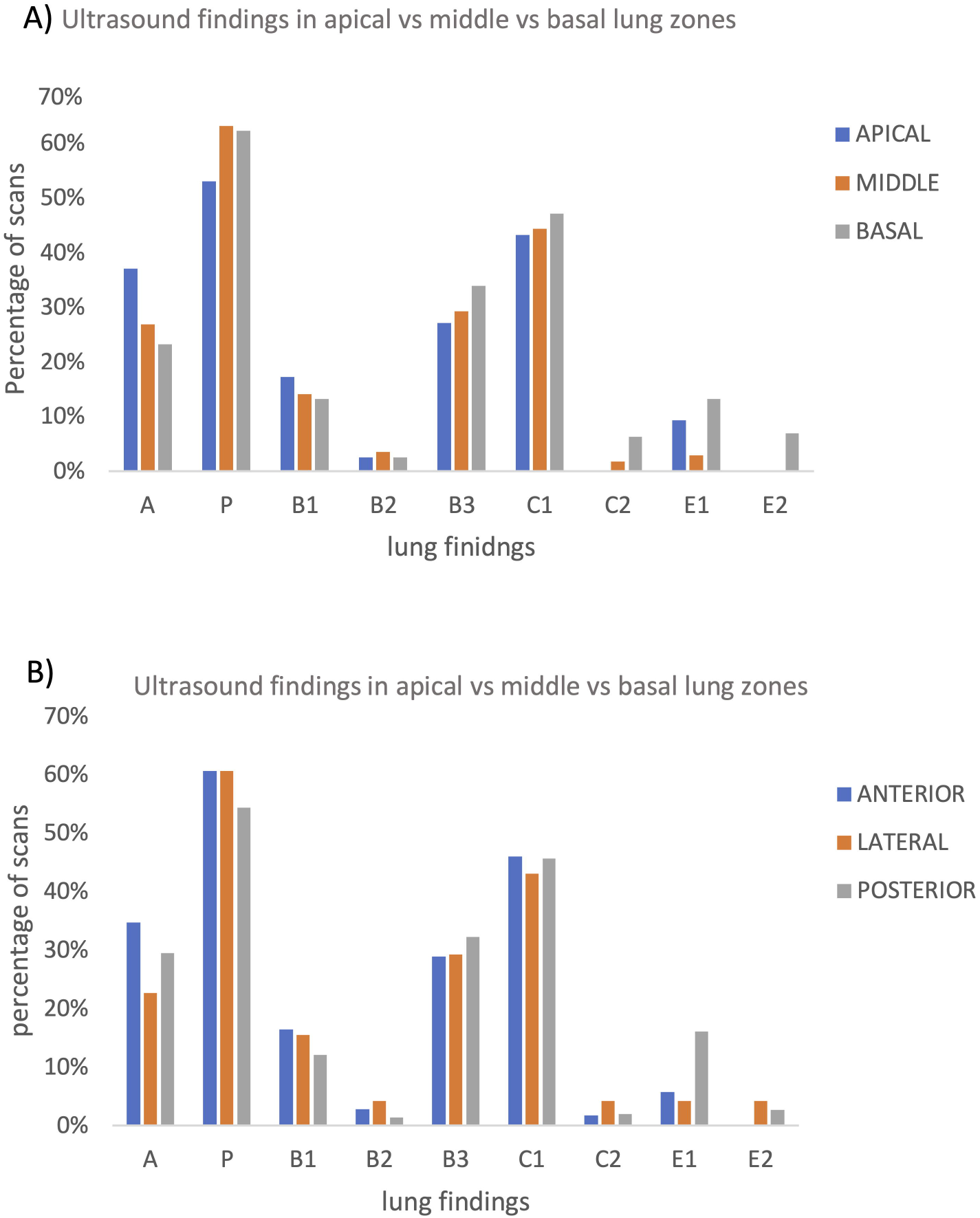
Bar Graphs depicting the frequency of abnormalities found in different areas of the lung. A) comparison of lung findings between transverse sections of the lung (apical, middle, basal), B) comparison of lung findings between coronal sections of the lung (anterior, lateral, posterior)

### Distribution of lung abnormalities amongst different clinical severity groups

The results of comparing lung findings between clinical severity groups can be found in Table 5. The percentage of A -lines was significantly higher in scans of patients with mild disease, occurring in 57.1% (C.I.95% 50.6-63.6%) compared to 11.8% (C.I.95% 6.4-17.2%), and 6.0% (C.I.95% 2.0-10.1%) in moderate and severe severity groups. Conversely, irregular pleura, subpleural consolidations and confluent B-lines were all found to a significantly higher extent in the moderate and severe categories than in scans from patients with mild disease. Whereas irregular pleura was demonstrated in 40.0% (C.I.95% 33.3-46.1%) of lung scans in mild disease, the frequency rose to 64.7% (C.I.95% 56.7-72.7%) and 85.7% (C.I.95% 79.8-91.7%) in moderate and severe oxygen deficiency. Small subpleural consolidations were found in only 27.7% (C.I.95% 21.8-33.5%) of mild disease scans, as opposed to 52.2% (C.I.95% 43.8-60.6%) in scans from the moderate disease category and 66.2% (C.I.95% 58.1-74.2%) of the severe disease group. Wide B-lines or confluent B-lines spanning entire rib spaces (B3) are seen in 15.6% (C.I.95% 10.9-20.4%) of mild disease scans, and more than doubled to 38.2% (C.I.95% 30.1-46.4%) in moderate and 45.1% (C.I.95% 36.7-53.6%) in severe disease (Figure 6A)

**Table 5.**
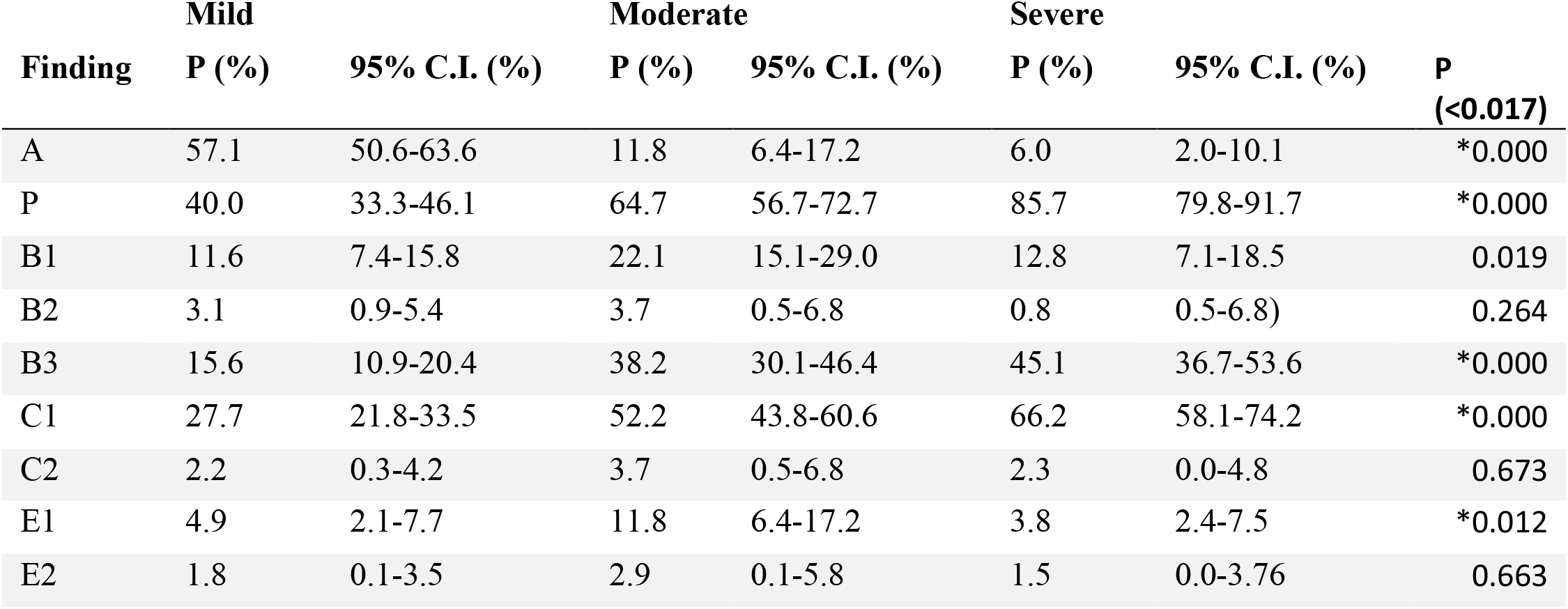
Frequency of lung abnormalities in different disease severities. The desired p-value was reached by Bonferroni correction to allow for multiple comparisons. Asteriks indicate statistical significance.

**Figure 6.**
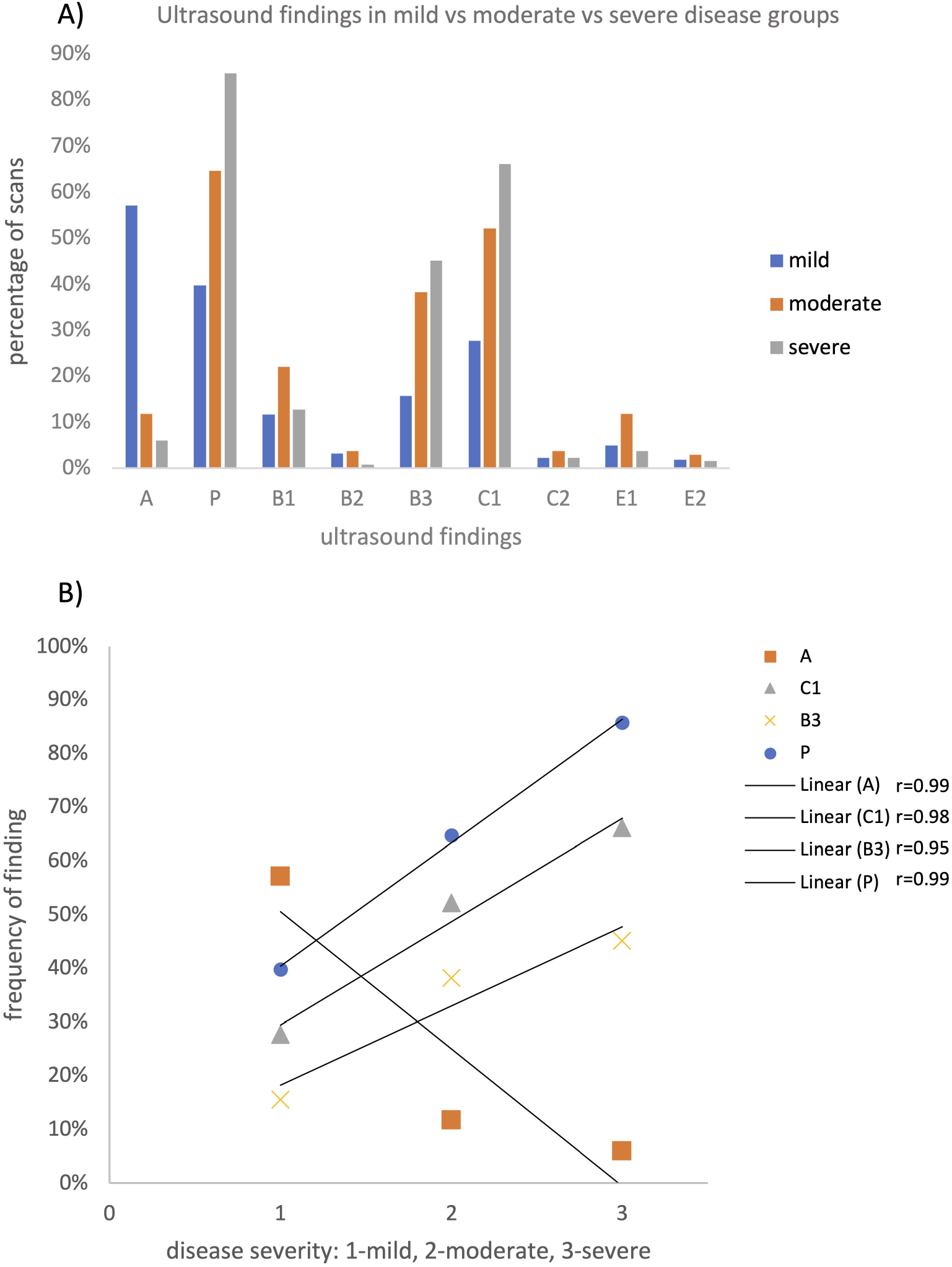
Comparisons of frequency of abnormalities found in different disease severity groups (mild, moderate, severe). A) Bar graph showing percentage of lung zones affected by each abnormality for each severity group, B) linear regression graph showing positive correlation between disease severity and A, P, C1 and B3 ultrasound patterns and negative correlation between A-lines and disease severity. Adjusted Pearson’s coefficient values (r) from linear regression analysis are indicated.

Interestingly, small peripleural effusions occurred in a significantly higher percentage of scans in the moderate disease group (11.8%, C.I.95% 6.4-17.2%) than in patients with mild (4.9% (C.I.95% 2.1-7.7%) or severe disease (3.8%, C.I.95% 2.4-7.5%).

Linear regression analysis showed a strong negative correlation between A-lines and clinical severity, with an adjusted Pearson’s correlation coefficient (r) of −0.99. A strong positive correlation was found between pleural irregularity and clinical severity (r = 0.99), and C1 and clinical severity (r = 0.98). B3 also displayed a positive correlation with clinical severity (r = 0.95) (Figure 6B).

## DISCUSSION

Our analysis of lung ultrasound scans of COVID-19 positive patients resulted in a clear set of abnormalities seen on lung ultrasound in the context of SARS-CoV-2 infection. These include pleural irregularities, small peripheral consolidations, a range of B-line patterns from separate thin B-lines to completely coalesced B-lines resulting in the appearance of a white intercostal space, as well as small peri-pleural effusions. Large effusions and consolidations were less prominently featured which reflects findings of other groups.

Pleural irregularities (P) and small peripheral subpleural consolidations (<1cm) (C) are the most prevalent lung abnormalities found in patients with SARS-CoV-2 infection. They occur in all severity groups of infected patients with no predilection for gravity dependent zones. Both of these abnormalities can be seen frequently in infected patients with normal oxygenation (mild disease group), but increase significantly in moderate and severe disease, where they occur 85% and 67% of lung zones respectively.

The inverse relationship found between the occurrence of A-lines and disease severity is logical, as the amount of interspersed normal lung, demonstrated by A-lines is replaced by abnormal lung in more severe disease.

Wide, thick B-lines moving with the pleura and occupying a large percentage of the pleural space have been described before in connection with COVID-19 respiratory disease and have been coined a variety of terms including ‘waterfall sign’ and ‘lightbeam sign’ (20)(15). Our findings echo these previous accounts. Wide B-lines that occupy >25% of the pleural space are seen in all disease severities, but whilst they are seen in about 15% of lung zones in mild disease, they are seen on more than double this frequency in moderate and further increase in frequency in severe disease. As disease severity increases, the density and width of these B-lines increases and can occupy the entire rib space, creating the appearance of a ‘white lung’. Although we could not see a statistically significant difference in anatomical distribution of wide B-lines, we saw a slight preference for posterolateral zones, particularly in milder disease. The presence of confluent B-lines, particularly white rib spaces in anterior zones appeared to indicate more severe disease. The small sample size in our study might be responsible for the lack of detecting a statistical significance.

B-lines indicate alveolar-interstitial abnormalities and are therefore seen in a variety of pathologies such as pneumonia, pulmonary oedema and interstitial lung disease. From research surrounding ARDS and pulmonary oedema, the amount of B-lines is linked to loss of aeration and has been used previously to develop a scoring system to predict disease severity and progression, assess therapeutic effect, or guide ventilation strategies(33)(34)(35). This appears to be similar in COVID-19 pneumonia, indicating these lung pathologies share important characteristics on lung ultrasound. In fact, this is also confirmed by appearances on CT scans, where alveolar-interstitial syndromes are seen as GGO on CT which is a pathognomonic finding in COVID-19 lung disease.

It has to be noted that we largely followed practises in the leading literature for the analysis of B-lines (36)(37)(38). We used simplified semiquantitative numerical evaluation of B-lines, where B-lines were counted per intercostal space and grouped into 3 categories (B1, B2, B3). To incorporate the frequent and new finding of wide B-lines into the scoring system, we scored them as B3 if they occupied >25% of the pleural space at origin. In this context, new consensus definitions may be required to score B-lines, which may help in differentiating different aetiologies of lung pathology from COVID-19.

Thin and scattered B-lines (B1) were incorporated in the analysis to elicit their significance in the context of COVID-19. In different context, it is known that <2 thin B-lines can be found in lung bases without signifying a disease process(36)(38). Our results did not challenge this notion with no difference in the B1 pattern seen between disease severities or areas of the lung.

Small (<1cm) localised effusions were a finding in our study which has been noted previously(20) and has been discussed in expert circles but has not attracted widespread attention in the literature. These small fluid collections appear to be most prevalent in moderate disease. The clinical significance of this finding is yet to be elicited but it is conceivable that it represents localised fluid collections as a result of inflammatory reactions.

Interestingly, when looking at the distribution of lung abnormalities within different areas of the lung, we could not find significant differences in distribution of lung findings commonly seen in COVID-19 respiratory disease. This is in agreement with the classic findings seen on CT describing peripheral GGO with spared areas occurring anywhere in the lung. A slight preference of abnormalities for posterobasal areas can be seen when gravity-dependent abnormalities such as larger consolidations and pleural effusions are also taken into account as has been done in previous studies(9)(20)(11).

With lung changes on ultrasound correlating to disease severity, a scoring system to predict disease severity or guide therapeutic approaches becomes conceivable. The ‘lung ultrasound score’ (LUS) described in the context of cardiogenic pulmonary oedema has been suggested (39) which converts numerically evaluated B-lines into a score to predict disease severity and progression, assess therapeutic effect, or guide ventilation strategies(33)(34)(35). The LUS scoring system however does not take into account pleural changes. Others have suggested to score COVID-19 changes on lung ultrasound based on pleural changes(40), but this approach neglects B-lines. We propose, that based on the results described here, any scoring system should negatively weight the presence of A-lines, positively weight pleural irregularities, small subpleural consolidations and broad B-lines.

It is important to remember that the lung changes seen ultrasound in COVID-19 patients are not specific to COVID-19 and with the decline in prevalence of this disease caution needs to be taken for potential over- and misdiagnosis. Other causes of viral pneumonia such as H1N1 (swine flu) or H7N9 (avian flu), have shown similar findings on lung ultrasound (8)(41), and further research is needed to investigate whether there are distinguishing factors between different types of viral pneumonias on lung ultrasound. In addition, little is known about the temporal progression of COVID-19 lung changes which is an important factor in deciding whether lung ultrasound is as useful for the future evaluation of SARS-CoV-2 infection as it was in the first ‘wave’.

There are several limitations to our study. The sample size was small but given the unique situation with a fast-evolving pandemic we analysed important lung ultrasound characteristics in the limited time available and used statistical measures to allow for small sample sizes. In addition, our most significant findings had non-overlapping confidence intervals which provides additional confidence in the validity of our findings.

A potential shortcoming is also our classification of disease severity, primarily that the Berlin criteria classify severity based on PaO_2_, measured at a PEEP of 5(29). As not all our patients with mild symptoms were subjected to arterial blood gas analysis, we refer to research conducted by Rice and colleagues which showed that S/F can be reliably related and converted to P/F ratio(31). This has also been acknowledged by the authors of the original Berlin criteria(32). We use the Berlin criteria merely as a guide to classify severity.

In summary, in this study we have systematically analysed the lung ultrasound findings typical for COVID-19 respiratory disease for their overall frequencies as well as anatomical prevalence within the lung. Pleural irregularities, small peripheral consolidations and wide B-lines occur in all severities of COVID-19 infection, but their frequency also directly correlates to disease severity as defined by oxygenation deficit. Small localised peripleural effusions are a feature in COVID-19 respiratory disease.

This information can add to existing data and help validate the use of lung ultrasound in the context of COVID-19 starting from triage, diagnosis to prognosis and formulation of treatment strategy. As COVID-19 pandemic progresses, it would be imperative to investigate how long the lung changes last and whether lung ultrasound could be useful in disease follow-up.

## Data Availability

all data is provided in the article

## DECLARATIONS

### Ethics Approval

This study was approved by the Health Research Authority (HRA) (no.286642).

### Consent for Publication

Not applicable

### Availability of data and materials

All relevant data is included in the manuscript, tables and figures. Additional data can be requested from the corresponding author of this submission.

### Competing Interests

None declared

### Funding

No funding was requested for this study

## Acknowledgements

none

## Author’s Information

No additional information

## Notes

### Competing Interest Statement

The authors have declared no competing interest.

### Clinical Trial

The study is not a clinical trial, but a retrospective observational study using investigations as part of routine assessment

### Funding Statement

No external funding was received for this study

### Author Declarations

IRAS 286015 HRA approved - REC reference: 20/HRA/3386

### Summary of Updates

figures and statistics updated. Result section updated

## REFERENCES

1. Organisation WH. No Title [Internet]. WHO Director-General’s opening remarks at the media briefing on COVID-19. 2020 [cited 2020 Jun 23]. Available from: https://www.who.int/dg/speeches/detail/who-director-general-s-opening-remarks-at-the-media-briefing-on-COVID-1911-march-2020

2. Huang C, Wang Y, Li X, Ren L, Zhao J, Hu Y, et al. Clinical features of patients infected with 2019 novel coronavirus in Wuhan, China. The Lancet. 2020;

3. Yoon SH, Lee KH, Kim JY, Lee YK, Ko H, Kim KH, et al. Chest radiographic and ct findings of the 2019 novel coronavirus disease (Covid-19): Analysis of nine patients treated in korea. Korean J Radiol. 2020;

4. Li Y, Xia L. Coronavirus disease 2019 (COVID-19): Role of chest CT in diagnosis and management. Am J Roentgenol. 2020;

5. Salehi S, Abedi A, Balakrishnan S, Gholamrezanezhad A. Coronavirus disease 2019 (COVID-19): A systematic review of imaging findings in 919 patients. American Journal of Roentgenology. 2020.

6. Wong HYF, Lam HYS, Fong AHT, Leung ST, Chin TWY, Lo CSY, et al. Frequency and Distribution of Chest Radiographic Findings in Patients Positive for COVID-19. Radiology. 2020;

7. Agarwal PP, Cinti S, Kazerooni EA. Chest radiographic and CT findings in novel swine-origin influenza A (H1N1) virus (S-OIV) infection. Am J Roentgenol. 2009;

8. Tsai NW, Ngai CW, Mok KL, Tsung JW. Lung ultrasound imaging in avian influenza A (H7N9) respiratory failure. Crit Ultrasound J. 2014;

9. Peng QY, Wang XT, Zhang LN. Findings of lung ultrasonography of novel corona virus pneumonia during the 2019–2020 epidemic. Intensive Care Medicine. 2020.

10. Vetrugno L, Bove T, Orso D, Barbariol F, Bassi F, Boero E, et al. Our Italian experience using lung ultrasound for identification, grading and serial follow-up of severity of lung involvement for management of patients with COVID-19. Echocardiography. 2020.

11. Volpicelli G, Gargani L. Sonographic signs and patterns of COVID-19 pneumonia. Ultrasound J. 2020;

12. Xing C, Li Q, Du H, Kang W, Lian J, Yuan L. Lung ultrasound findings in patients with COVID-19 pneumonia. Crit Care. 2020;

13. Yasukawa K, Minami T. Point-of-Care Lung Ultrasound Findings in Patients with Novel Coronavirus Disease (COVID-19) Pneumonia. Am J Trop Med Hyg. 2020;

14. Lomoro P, Verde F, Zerboni F, Simonetti I, Borghi C, Fachinetti C, et al. COVID-19 pneumonia manifestations at the admission on chest ultrasound, radiographs, and CT: single-center study and comprehensive radiologic literature review. Eur J Radiol Open. 2020;

15. Volpicelli G, Lamorte A, Villén T. What’s new in lung ultrasound during the COVID-19 pandemic. Intensive Care Med. 2020;

16. Castelao J, Graziani D, Soriano JB, Izquierdo JL. Findings and Prognostic Value of Lung Ultrasound in COVID-19 Pneumonia. J Ultrasound Med. 2020;

17. Colombi D, Petrini M, Maffi G, Villani GD, Bodini FC, Morelli N, et al. Comparison of admission chest computed tomography and lung ultrasound performance for diagnosis of COVID-19 pneumonia in populations with different disease prevalence. Eur J Radiol. 2020;

18. Narinx N, Smismans A, Symons R, Frans J, Demeyere A, Gillis M. Feasibility of using point-of-care lung ultrasound for early triage of COVID-19 patients in the emergency room. Emerg Radiol. 2020;

19. Karagöz A, Sağlam C, Demirbaş HB, Korkut S, Ünlüer EE. Accuracy of Bedside Lung Ultrasound as a Rapid Triage Tool for Suspected Covid-19 Cases. Ultrasound Q. 2020;

20. Huang Y, Wang S, Liu Y, Zhang Y, Zheng C, Zheng Y, et al. A Preliminary Study on the Ultrasonic Manifestations of Peripulmonary Lesions of Non-Critical Novel Coronavirus Pneumonia (COVID-19). SSRN Electron J. 2020;

21. Zhou P, Yang X Lou, Wang XG, Hu B, Zhang L, Zhang W, et al. A pneumonia outbreak associated with a new coronavirus of probable bat origin. Nature. 2020;

22. Lu W, Zhang S, Chen B, Chen J, Xian J, Lin Y, et al. A Clinical Study of Noninvasive Assessment of Lung Lesions in Patients with Coronavirus Disease-19 (COVID-19) by Bedside Ultrasound. Ultraschall Med. 2020;

23. Poggiali E, Dacrema A, Bastoni D, Tinelli V, Demichele E, Ramos PM, et al. Can lung US Help critical care clinicians in the early diagnosis of novel coronavirus (COVID-19) pneumonia? Radiology. 2020.

24. G. T, X. L, Z. Z, Z. W, F. H, Y. Z, et al. Use of Lung Ultrasound to Differentiate Coronavirus Disease 2019 (COVID-19) Pneumonia From Community-Acquired Pneumonia. Ultrasound Med Biol. 2020;

25. Lichter Y, Topilsky Y, Taieb P, Banai A, Hochstadt A, Merdler I, et al. Lung ultrasound predicts clinical course and outcomes in COVID-19 patients. Intensive Care Med. 2020;

26. Zhang Y, Xue H, Wang M, He N, Lv Z, Cui L. Lung Ultrasound Findings in Patients With Coronavirus Disease (COVID-19). Am J Roentgenol. 2020;

27. Mohamed MFH, Al-Shokri S, Yousaf Z, Danjuma M, Parambil J, Mohamed S, et al. Frequency of abnormalities detected by point-of-care lung ultrasound in symptomatic COVID-19 patients: Systematic review and meta-analysis. American Journal of Tropical Medicine and Hygiene. 2020.

28. Intensive Care Society. National Service Evaluation of Lung and Heart Ultrasound in Patients with Suspected or Proven COVID Project [Internet]. 2020 [cited 2020 Oct 25]. Available from: https://members.ics.ac.uk/ICS/ICS/FUSIC/FUSIC_COVID-19.aspx

29. The ARDS Definition Task Force*. Acute Respiratory Distress Syndrome<subtitle>The Berlin Definition</subtitle><alt-title>The Berlin Definition of ARDS</alt-title>. JAMA J Am Med Assoc. 2012;

30. No Title. Coronavirus (COVID-19): guidance. 2020.

31. Rice TW, Wheeler AP, Bernard GR, Hayden DL, Schoenfeld DA, Ware LB. Comparison of the SpO2/FIO2 ratio and the PaO 2/FIO2 ratio in patients with acute lung injury or ARDS. Chest. 2007;

32. Ferguson ND, Fan E, Camporota L, Antonelli M, Anzueto A, Beale R, et al. The Berlin definition of ARDS: An expanded rationale, justification, and supplementary material. Intensive Care Med. 2012;

33. Bello G, Blanco P. Lung ultrasonography for assessing lung aeration in acute respiratory distress syndrome: A narrative review. Journal of Ultrasound in Medicine. 2019.

34. Bouhemad B, Mongodi S, Via G, Rouquette I. Ultrasound for ‘lung monitoring’ of ventilated patients. Anesthesiology. 2015.

35. Chiumello D, Mongodi S, Algieri I, LucaVergani G, Orlando A, Via G, et al. Assessment of lung aeration and recruitment by CT scan and ultrasound in acute respiratory distress syndrome patients. Crit Care Med. 2018;

36. Lichtenstein D, Mézière G, Biderman P, Gepner A, Barré O. The comet-tail artifact: An ultrasound sign of alveolar-interstitial syndrome. Am J Respir Crit Care Med. 1997;

37. Lichtenstein DA. Lung ultrasound in the critically ill: The blue protocol. Lung Ultrasound in the Critically Ill: The BLUE Protocol. 2015.

38. Lichtenstein DA. Current Misconceptions in Lung Ultrasound: A Short Guide for Experts. Chest. 2019;

39. Soummer A, Perbet S, Brisson H, Arbelot C, Constantin JM, Lu Q, et al. Ultrasound assessment of lung aeration loss during a successful weaning trial predicts postextubation distress. Crit Care Med. 2012;

40. Soldati G, Smargiassi A, Inchingolo R, Buonsenso D, Perrone T, Briganti DF, et al. Proposal for International Standardization of the Use of Lung Ultrasound for Patients With COVID-19: A Simple, Quantitative, Reproducible Method. J Ultrasound Med. 2020;

41. Testa A, Soldati G, Copetti R, Giannuzzi R, Portale G, Gentiloni-Silveri N. Early recognition of the 2009 pandemic influenza A (H1N1) pneumonia by chest ultrasound. Crit Care. 2012;

